# Genomic surveillance of multidrug-resistant organisms based on long-read sequencing

**DOI:** 10.1101/2024.02.18.24301916

**Authors:** Fabian Landman, Casper Jamin, Angela de Haan, Sandra Witteveen, Jeroen Bos, Han G.J. van der Heide, Leo M. Schouls, Antoni P.A. Hendrickx, Dutch CPE and MRSA surveillance study groups

## Abstract

**Background:** Multidrug-resistant organisms (MDRO) pose a significant threat to public-health world-wide. The ability to identify antimicrobial resistance determinants, to assess changes in molecular types, and to detect transmission are essential for effective surveillance and infection prevention of MDRO. Molecular characterization based on long-read sequencing has emerged as a promising alternative to short-read sequencing. The aim of this study was to rapidly and accurately characterize MDRO for surveillance and transmission studies based on long-read sequencing only.

**Methods:** Genomic DNA of 356 MDRO was automatically extracted using the Maxwell-RSC48. The MDRO included 106 *Klebsiella pneumoniae* isolates, 85 *Escherichia coli*, 15 *Enterobacter cloacae* complex, 10 *Citrobacter freundii*, 34 *Pseudomonas aeruginosa*, 16 *Acinetobacter baumannii*, and 69 methicillin-resistant *Staphylococcus aureus* (MRSA), of which 24 were from an outbreak. MDRO were sequenced using both short-read (Illumina NextSeq 550) and long-read (Nanopore Rapid Barcoding Kit-24-V14, R10.4.1) whole-genome sequencing (WGS). Basecalling was performed for two distinct models using Dorado-0.3.2 duplex mode. Long-read data was assembled using Flye, Canu, Miniasm, Unicycler, Necat, Raven and Redbean assemblers. Long-read WGS data with >40x coverage was used for multi-locus sequence typing (MLST), whole-genome MLST (wgMLST), *in silico* multiple locus variable-number of tandem repeat analysis (iMLVA) for MRSA, and identification of resistance genes (Abricate).

**Results:** Comparison of wgMLST profiles based on long-read and short-read WGS data revealed >95% of wgMLST profiles within the species-specific cluster cut-off, except for *P. aeruginosa.* The wgMLST profiles obtained by long-read and short-read WGS differed only one to nine wgMLST alleles for *K. pneumoniae*, *E. coli*, *E. cloacae* complex, *C. freundii*, *A. baumannii* complex and MRSA. For *P. aeruginosa* differences were up to 27 wgMLST alleles between long-read and short-read wgMLST. MLST sequence types and *in silico* MLVA types were concordant between long-read and short-read WGS data and conventional MLVA typing. Antimicrobial resistance genes were detected in long-read sequencing data with high sensitivity/specificity (92-100%/99-100%). Long-read sequencing enabled analysis of an MRSA outbreak.

**Conclusions:** We demonstrate that molecular characterization of automatically extracted DNA followed by long-read sequencing is as accurate and more cost-effective compared to short-read sequencing. Long-read sequencing is suitable for molecular typing and outbreak analysis as part of genomic surveillance of MDRO. However, the analysis of *P. aeruginosa* requires further improvement which may be obtained by other basecalling algorithms. The low implementation costs, low price per isolate, and rapid library preparation for long-read sequencing of MDRO extends its applicability to resource-constrained settings and low-income countries world-wide.

## Introduction

Multidrug-resistant organisms (MDRO) such as methicillin-resistant *Staphylococcus aureus* (MRSA), carbapenemase-producing *Enterobacterales* (CPE) and *Pseudomonas aeruginosa* (CPPA) and carbapenem-resistant *Acinetobacter baumannii* (CRAB) pose a growing public health concern worldwide. The ability to type MDRO accurately and rapidly and to identify resistance determinants is essential for effective surveillance and infection control. National reference laboratories and medical microbiology laboratories historically typed MDRO using various classical typing methods such as phage typing [1], pulsed-field gel electrophoresis (PFGE) [2,3], amplified fragment length polymorphism (AFLP) [4], Staphylococcal protein A (Spa)-typing [5,6], multiple locus variable-number of tandem repeat analysis (MLVA), but also DNA sequencing-based typing methods such as multi-locus sequence typing (MLST) [7–10]. These methods are being phased out by whole-genome sequencing (WGS)-based methods such as core genome (cg) or whole genome (wg) MLST, as these methods significantly increased typing resolution. In addition, WGS data enable identification of resistance and virulence genes [11]. Current WGS-based methods typically use short-read sequencing technologies with read lengths of 150 bases, such as Illumina next-generation sequencing, and have been widely used for public health genomic surveillance, molecular typing, and outbreak detection of MDRO [12,13]. Short-read WGS costs are high and force concessions for national and local surveillance to sequence only a limited selection of MDRO [14]. Additionally, short-read sequencing technologies have limitations in detecting structural variations, such as large insertions, deletions, inversions, repetitive sequence elements and antimicrobial resistance (AMR) plasmids, all common in bacterial genomes [13]. Reconstruction of complete bacterial genomes by *de novo* assembly is unattainable through short-read sequencing as these structural variations and genomic elements are larger than individual reads. Thus, chromosomes and AMR encoding plasmids fail to assemble into complete assemblies. Hybrid assemblies of short-read and long-read WGS data enabled reconstruction of MDRO genomes but is laborious and more expensive than short-read sequencing alone. Long-read sequencing technologies, such as Nanopore sequencing, can overcome these problems and have recently emerged as a promising alternative to short-read sequencing or hybrid assemblies [15–18]. Furthermore, the accuracy of Nanopore long-read sequencing significantly improved over time (https://nanoporetech.com/platform/accuracy). The major objective of this study was to determine whether long-read sequencing can replace short-read sequencing to enable classical and new typing methods such as MLVA, MLST and wgMLST in the genomic surveillance and outbreak analysis of MDRO.

## Methods

### Multidrug-resistant organisms (MDRO) from the Dutch national surveillance 2023

Medical microbiology laboratories (MML) in the Netherlands are requested to submit *Enterobacterales, P. aeruginosa, A. baumannii* suspected for carbapenemase production or carbapenem resistance, and MRSA isolates cultured for patient care (from symptomatic infections or asymptomatic carriership), to the National Institute for Public Health and the Environment (RIVM) [14,19]. Between 1 May and 30 September 2023, 3,138 suspected MDRO were received by the RIVM of which the majority 2,235 were MRSA isolates. lllumina short-read sequencing and Nanopore long-read sequencing were performed on 356 MDRO in this study (*vide infra*). The MDRO included 106 *Klebsiella pneumoniae* isolates, 85 *Escherichia coli*, 15 *Enterobacter cloacae* complex, 10 *Citrobacter freundii*, 34 *Pseudomonas aeruginosa*, 16 *Acinetobacter baumannii*, 69 methicillin-resistant *Staphylococcus aureus* (MRSA) and other species (Table 1, Supplementary Table 1).

**Table 1.** Selected MDRO from the Dutch national surveillance May-September 2023.

| **MDRO** | **All coverages** | **>20x coverage** | **>30x coverage** | **>40x coverage** |
| --- | --- | --- | --- | --- |
| *Acinetobacter baumannii* | 16 | 11 | 7 | 4 |
| *Acinetobacter nosocomialis* | 1 | 1 | 1 | 1 |
| *Acinetobacter ursingii* | 1 |  |  |  |
| *Citrobacter farmeri* | 1 | 1 | 1 | 1 |
| *Citrobacter freundii* | 10 | 9 | 9 | 9 |
| *Enterobacter cloacae complex* | 15 | 14 | 11 | 11 |
| *Escherichia coli* | 85 | 77 | 69 | 54 |
| *Klebsiella oxytoca* | 6 | 5 | 3 | 2 |
| *Klebsiella pneumoniae* | 106 | 84 | 70 | 48 |
| *Proteus mirabilis* | 4 | 4 | 4 | 2 |
| *Providencia stuartii* | 3 | 2 | 2 | 2 |
| *Pseudomonas aeruginosa* | 34 | 23 | 12 | 7 |
| *Pseudomonas nitroreducens* | 1 |  |  |  |
| *Raoultella ornithinolytica* | 4 | 2 | 1 |  |
| *Staphylococcus aureus* | 45 | 45 | 44 | 39 |
| *Staphylococcus aureus* (outbreak) | 24 | 23 | 19 | 16 |
| **Total** | **356** | **301** | **253** | **196** |

### Automated Oxford Nanopore Technologies long-read sequencing

The genomic DNA of MDRO was isolated using The Maxwell® (Promega), an automated nucleic acid extraction and purification platform. The Maxwell RSC Cultured Cells DNA kit (AS1620) kit was used to isolate up to 48 bacterial isolates per run. Manufacturer’s instructions were followed, except using nuclease-free water instead of TE buffer to create the cell suspension and without RNase treatment. The Oxford Nanopore protocol for rapid sequencing DNA V14 – barcoding SQK-RBK114.24 was used (Oxford Nanopore Technologies [ONT], https://nanoporetech.com/). In brief, barcoded transposome complexes were used to tagment the DNA and simultaneous attachment of a pair of barcodes. Routine isolations had shown DNA concentrations within 50-300 ng/μl, thus, for maintaining high throughput capacity, DNA concentrations were not measured prior to library preparation. All samples were pooled and after clean-up the sequencing adapters (RAP reagent) were added. Sequencing buffer and library beads were added, and the final library was loaded onto a MinION flow cell (FLO-MIN114, R10.4.1) (https://community.nanoporetech.com/docs/prepare/library_prep_protocols/rapid-sequencing-gdna-barcoding-sqk-rbk114/v/rbk_9176_v114_revm_27nov2022). The 48-h sequence run was started with live basecalling and demultiplexing enabled using the MinKNOW software through a GridION device with 5 kHz data acquisition enabled for all samples. However, the actual basecalling for analysis in this study was performed on the resulting barcoded pod5 directories using Dorado 0.3.2 duplex mode with the optional flags --guard-gpus, -b 448 and writing data to Fastq format. We assessed the performance of two basecalling models, dna_r10.4.1_e8.2_400bps_sup@v4.2.0 and res_dna_r10.4.1_e8.2_400bps_sup@2023-09-22_bacterial-methylation. These models are referred to as Dorado (https://github.com/nanoporetech/dorado) and Rerio (https://github.com/nanoporetech/rerio), respectively. Data was assembled using an in-house Snakemake workflow. All bioinformatic tools were used with default parameters unless specified otherwise. First, Chopper v0.6.0 is used to extract all Q12 reads >1000-bp [20]. Additionally, 80-bp is cropped from both sides to remove possible adapters. Subsequently, FiltLong 0.2.1 was used to keep the 90% best scoring reads based on length and identity (https://github.com/rrwick/Filtlong). To assess the performance of different *de novo* assemblers long-read data was assembled using Canu v2.2 [21], Flye v2.9.2 [22], Minimap v2 2.26 followed by Miniasm v0.3 [23], Minipolish v0.1.3, Necat v0.0.1 [24], Raven v1.8.1 [25], Redbean wtdbg2.5 [26], and Unicycler v0.5.0 [27] long-read option. Furthermore, genome assembly polishing with Medaka v1.8.0 (https://github.com/nanoporetech/medaka) was used to test if this improved *de novo* assemblies, compared to unpolished assemblies. Medaka polishing was only done for Dorado basecalled reads, as there was no Medaka polishing model for Rerio basecalled data available during this study.

### Automated Illumina short-read sequencing

For in-house Illumina short-read sequencing of MDRO, the same DNA isolation method as described above was followed. DNA libraries were prepared using Nextera DNA Flex Library Prep kit (Illumina, San Diego, USA), followed by paired-end sequencing (2 x 150-bases) on the Illumina NextSeq550 platform (Illumina, USA), according to the manufacturer’s instructions. Read quality analysis and *de novo* assembly was performed with the Juno-assembly v2.0.2 pipeline (https://github.com/RIVM-bioinformatics/juno-assembly). Briefly, read quality assessment and filtering were done using FastQC and FastIP [28,29]. Genomes were assembled using SPAdes [30] and curated with QUAST [31], CheckM [32] and Bbtools (https://sourceforge.net/projects/bbmap/).

### Molecular typing from long-read and short-read sequencing data and data analyses

Sixty-nine MRSA isolates, including 24 outbreak isolates were characterized by multiple-locus variable number of tandem repeat analysis (MLVA) *in vitro* at BaseClear (Leiden, the Netherlands) and simultaneous PCR detection of the *mecA/mecC,* and *lukF*-PV/*lukS*-PV genes, the latter indicative of PVL production, as previously described [33,34]. In addition, *in silico* (i)MLVA on MRSA isolates was done in the RIVM on long-read WGS data only. The identification of resistance genes and replicons of MDRO was performed using ResFinder (version 4.1.11) and PlasmidFinder (version 2.1) databases from the Center for Genomic Epidemiology, using ABRicate (v1.0.1, https://github.com/tseemann/abricate). The resulting FASTA files were subjected to their corresponding species-specific multi-locus sequence typing (MLST) and whole-genome MLST (wgMLST) schemes using Ridom software in SeqSphere v9.0.1 if available [35]. The following in-house wgMLST schemes were used for analyses: the *A. baumannii* wgMLST scheme was based on 3,473 genes (2,390 core-genome genes and 1,083 accessory-genome genes, cut-off 15 wgMLST alleles), using *A. baumannii* strain ACICU (Genbank accession number NC_010611.1, September 2021) as a reference genome. The *K. pneumoniae* wgMLST scheme comprised 4,978 genes (3,471 core-genome and 1,507 accessory-genome targets, cut-off 20 wgMLST alleles) using *K. pneumoniae* MGH 78,578 (NC_009648.1) as a reference genome [19]. The *E. coli* wgMLST scheme comprised 4,503 genes (3,199 core-genome and 1,304 accessory-genome targets, cut-off 25 wgMLST alleles) using *E. coli* 536 (CP000247.1) as a reference genome [19]. The *P. stuartii* scheme comprised 3,744 genes (3,079 core-genome and 665 accessory-genome targets, cut-off 15 wgMLST alleles) with *P. stuartii* CP014024.2 as reference genome (Witteveen *et al.*, 2024 submitted). The *P. aeruginosa* wgMLST scheme included 6,442 genes (6,117 core-genome and 325 accessory-genome targets, cut-off 15 wgMLST alleles) using *P. aeruginosa* PAO1 (NC_002516.2) as a reference genome. The *P. mirabilis* wgMLST scheme included 3,517 genes (2,675 core-genome and 842 accessory-genome targets, cut-off 15 wgMLST alleles) using *P. mirabilis* HI4320 (NZ_CP042907.1) as a reference genome. The *C. freundii* wgMLST scheme included 4,495 genes (2964 core-genome and 1531 842 accessory-genome targets, cut-off 20 wgMLST alleles) using *C. freundii* strain HM38 (CP024672.1) as a reference. For MRSA, the COL-based wgMLST scheme comprised 2,567 genes (1,861 core-genome and 706 accessory-genome targets, cut-off 15 wgMLST alleles) was used [14,36]. For *E. cloacae* complex a pgMLST scheme comprised of 9,829 genes from references CP001918, CP017186 and CP017184 was used, cut-off 20 wgMLST alleles [37]. Both MLST and wgMLST profiles were imported into BioNumerics version 8.1.1 (Applied Maths, Sint-Martens-Latem, Belgium) and used in cluster analyses. Missing data were ignored in the analyses. The Illumina short-read sequencing results were considered golden standard method for MLST sequence type determination, wgMLST allele calling, AMR gene and replicon identification, and long-read assembly methods throughout this study. All further data analyses and visualization was done using Python (v3.11.5, https://www.python.org/downloads/release/python-3115/), Pandas (v2.0.3, https://pandas.pydata.org/docs/index.html) and Plotly (v5.18.0, https://github.com/plotly/plotly.py).

### Data availability

Raw short-read and long-read sequencing data of 356 Dutch CPE/CPPA/CRAB/MRSA surveillance isolates have been deposited in the SRA database under BioProject numbers PRJNA1076692, PRJNA1076808, and PRJNA903550 (Supplementary Table 1). Relevant code was made available through https://github.com/RIVM-bioinformatics/in-silico-mlva. The authors confirm that all supporting data, code, protocols and accession numbers have been provided within the article and through supplementary data files.

## Results

### Assessing sequencing coverage and basecalling model for wgMLST on long-read sequenced MDRO

From May to September 2023, the National Institute for Public Health and the Environment (RIVM) received 3,138 MDRO. In this study, 356 genetically highly diverse MDRO were included; 269 CPE, of which the majority were *K. pneumoniae* (*n*=106) and *E. coli* (*n*=85) and nine other species (n=78), CPPA (*n*=34), CRAB (*n*=17) and MRSA (*n*=69) (Table 1, Supplementary Table 1). We also included 24 PVL-negative MRSA isolates belonging to a recurring fusidic acid-resistant impetigo-associated MRSA outbreak in the Netherlands [38]. We isolated genomic DNA using the Maxwell and sequenced 356 MDRO using both Nanopore and Illumina WGS platforms. Sequencing coverage for long-read sequencing was assessed from 356 MDRO to determine a suitable cut-off for sequencing depth. The median relative number of wgMLST alleles (Nanopore versus Illumina assemblies) were 0.45, 0.93, 0.99 for coverages up to 0-10x, 10-20x and 20-30x, respectively (Figure 1A). The number of wgMLST alleles identified remained stable at 1.00 with higher sequencing depth, indicating that the same number of alleles were identified in the Nanopore assemblies compared to the Illumina assemblies. This trend was seen across all species tested in this study (data not shown). The median number of different wgMLST alleles compared to the Illumina assemblies was 97, 45, 17, 10, 7, 4, 5 for coverages up to 0-10x, 10-20x, 20-30x, 30-40x, 40-50x, 50-60x, 60-70x, respectively (Figure 1B). The median distance with Illumina assemblies was relatively stable, between 4 to 7 wgMLST alleles, with coverages from 40x and higher. At a coverage 100x several outliers are observed, with differences up to 300 alleles. These outliers belonged all to one single *P. aeruginosa* isolate basecalled with Dorado and assembled with the different *de novo* assemblers used in this study. The 196 long-read sequenced MDRO with a coverage higher than 40x were used for further analyses. To determine which long-read sequencing data basecalling model performed best and if Medaka polishing (for Dorado only) improved assembly when comparing long-read assemblies to the Illumina short-read sequencing golden standard, we determined the number of wgMLST alleles difference for 196 MDRO using these methods (Figure 2). Across all species, Rerio performed best and had the lowest number of faulty wgMLST allele calls. The median number of wgMLST alleles difference for Dorado duplex, Dorado duplex with Medaka, and Rerio duplex was 2, 2, 2 (*A. baumannii*), 5.5, 3, 1 (*C. freundii*), 2, 1, 1 (*E. cloacae* complex), 6, 3, 3, (*E. coli)*, 23, 19.5, 9 (*K. pneumoniae)*, 120, 79, 26.5 (*P. aeruginosa*), and 4, 2, 2 (*S. aureus*), respectively. Rerio was able to improve the long-read sequencing data in such a way that the number of faulty wgMLST allele calls dropped from 87-148 (depending on the assembler) to merely 0-5 faulty alleles for one particular *C. freundii* isolate. Therefore, Rerio was used for further analyses.

**Figure 1.**
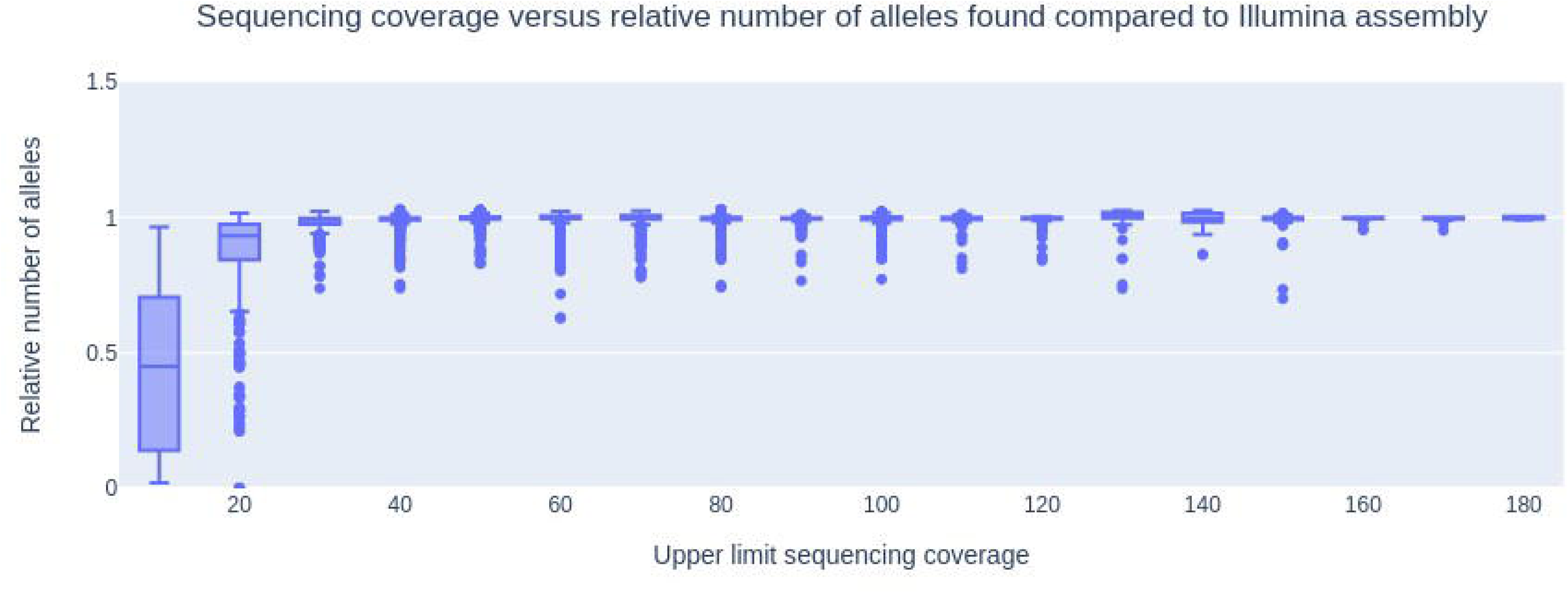
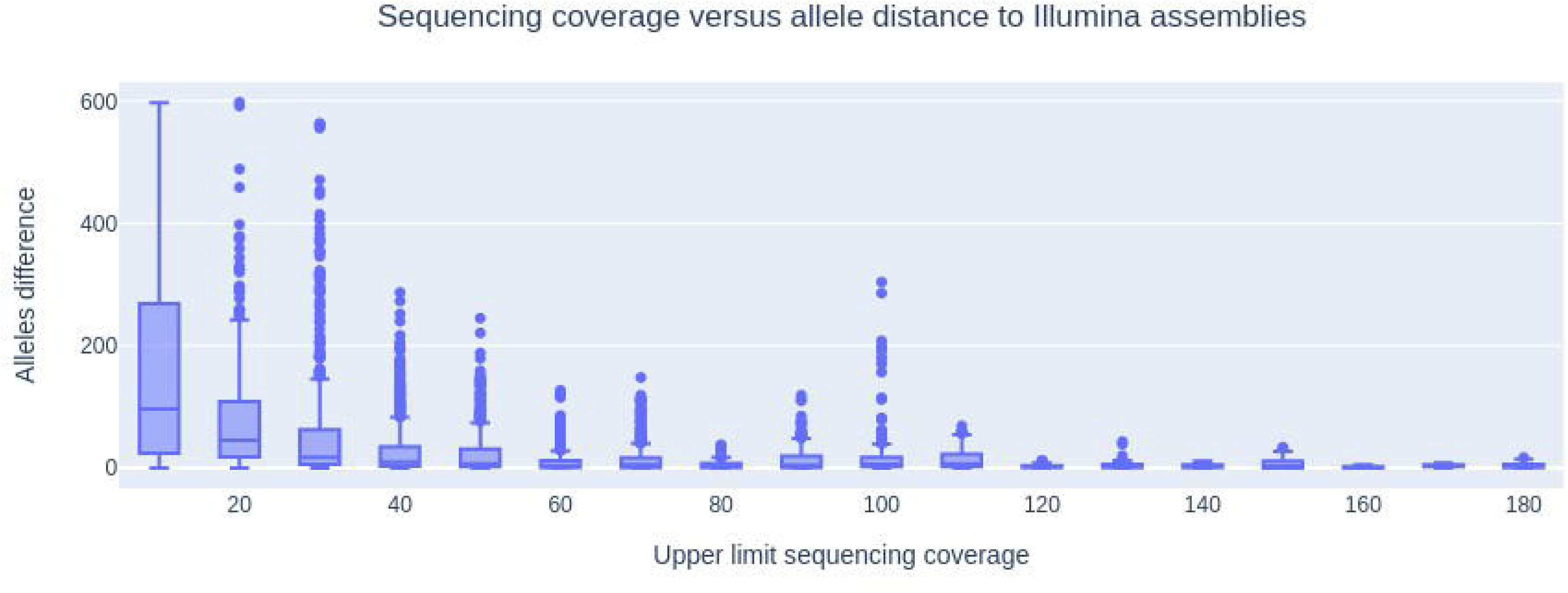
A) Nanopore long-read sequencing coverage versus relative number of wgMLST alleles identified in each assembly versus the Illumina genome assembly. B) Nanopore sequencing coverage versus wgMLST allele distance to Illumina assemblies.

**Figure 2.**
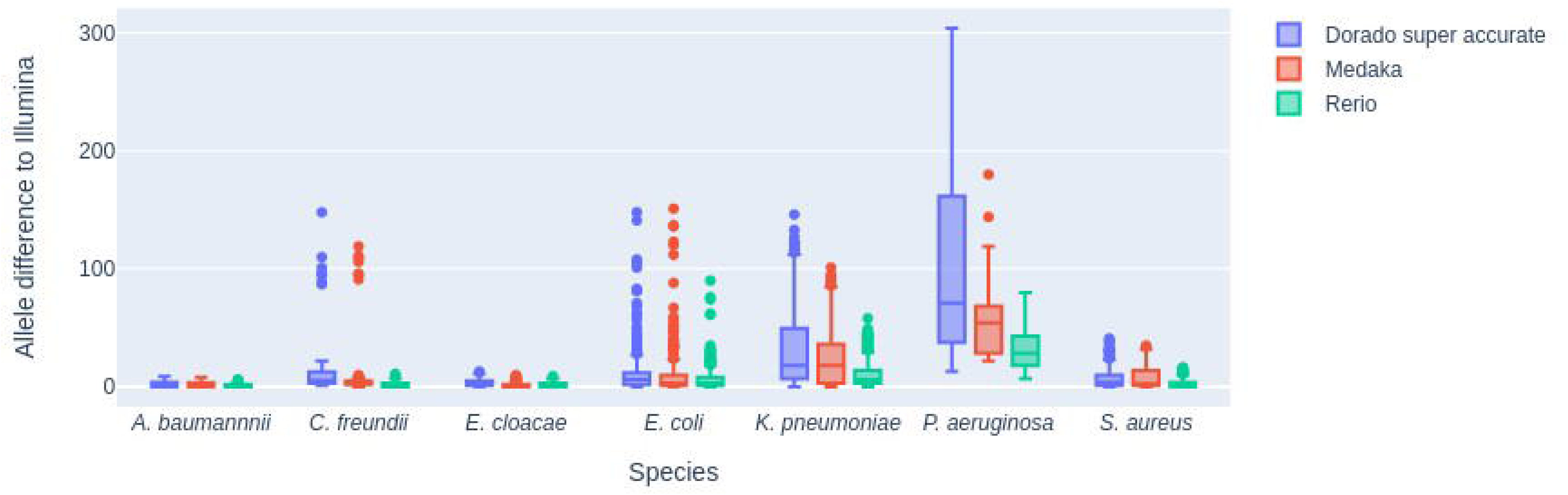
Nanopore long-read sequencing wgMLST alleles difference compared to Illumina assemblies, for Dorado super accurate, Rerio, and Medaka polishing method.

### Comparison of long-read assemblers for MLST and wgMLST on long-read sequenced MDRO

To assess the best assembler for *de novo* assembly of long-read only data in the absence of Illumina short-read sequencing data, Longcycler (Unicycler with long-reads only), Miniasm, Raven, Necat, Canu, Flye and Redbean were compared based on the median (50 percentile) wgMLST allele difference and the 95 percentile wgMLST alleles difference. With all species combined, the difference was 1; 18 (Canu), 1; 21.55 (Flye), 2.5; 15.55 (Longcycler), 2; 17 (Miniasm), 7; 35.55 (Necat), 3; 34.65 (Raven) and 6; 38.65 (Redbean) for the median and 95% percentile, respectively (Figure 3, Table 2). Although the median was slightly lower for Miniasm compared to Longcycler, the 95% performed better for Longcycler, and was therefore used for subsequent analyses. For each MDRO, the median and 95% percentile allele difference for Longcycler was 1; 2.0 (*A. baumannii*), 1; 4 (*C. freundii*), 1; 3.5 (*E. cloacae*), 2; 17 (*E. coli*), 4; 12.6 (*K. pneumoniae*), 2; 6 (*S. aureus*), and 25.5; 43 (*P. aeruginosa*), respectively (Table 2). Long-read sequencing-derived MLST sequence types (ST) were compared with short-read sequencing derived MLST STs among the MDRO tested. A few differences were observed, for two *E. coli* isolates the Illumina MLST ST did not call the *adk* gene, which was present in all other *E. coli* isolates and the long-read MLST STs all called the same locus. All other differences resulted from the long-read derived MLST missing a call compared to the Illumina MLST ST. For three out of four *A. baumannii* the *gdhB* gene was not called. Interruption by IS elements in this element has been previously noted and could explain its absence in all of the long-read MLST STs [39,40]. For Miniasm only one gene was not called in *K. pneumoniae* but otherwise was in concordance with all Illumina MLST STs and thus performed best. Longcycler also missed one additional gene in an *E. coli* isolate, but all other MLST STs were identical to the Illumina MLST STs (Supplementary Table 2).

**Figure 3.**
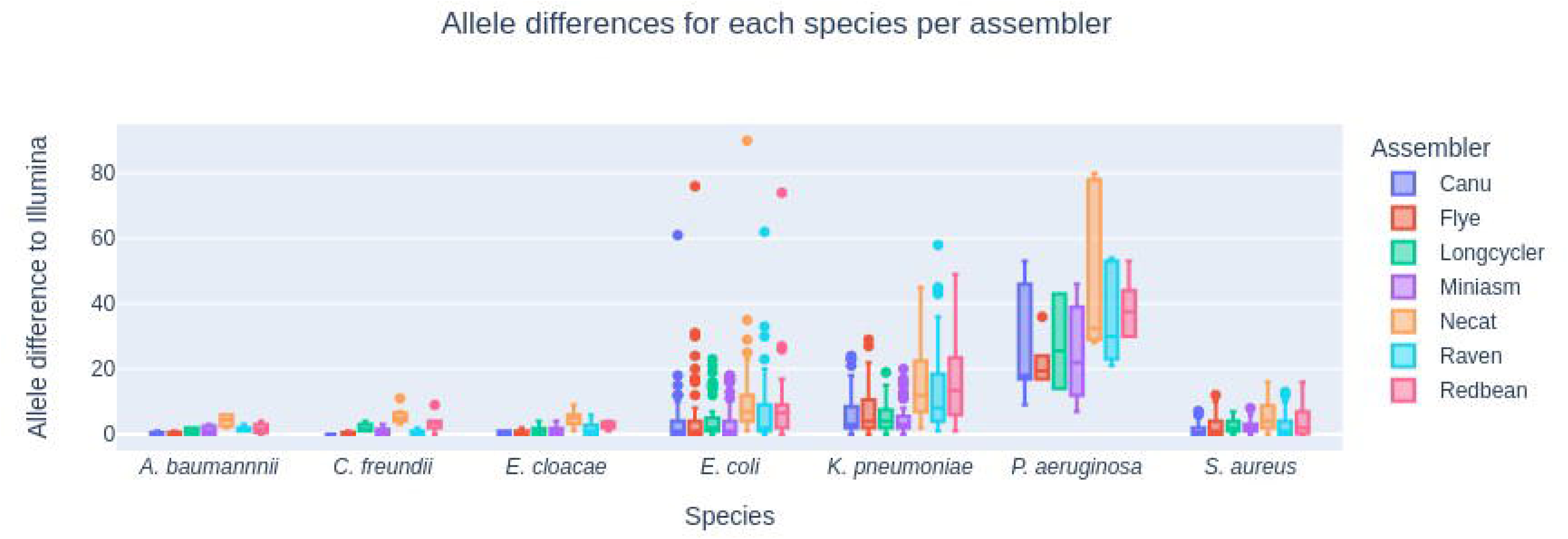
Nanopore long-read sequencing wgMLST alleles difference relative to Illumina short-read sequences per bacterial species and long-read sequence data assembler.

**Table 2.**
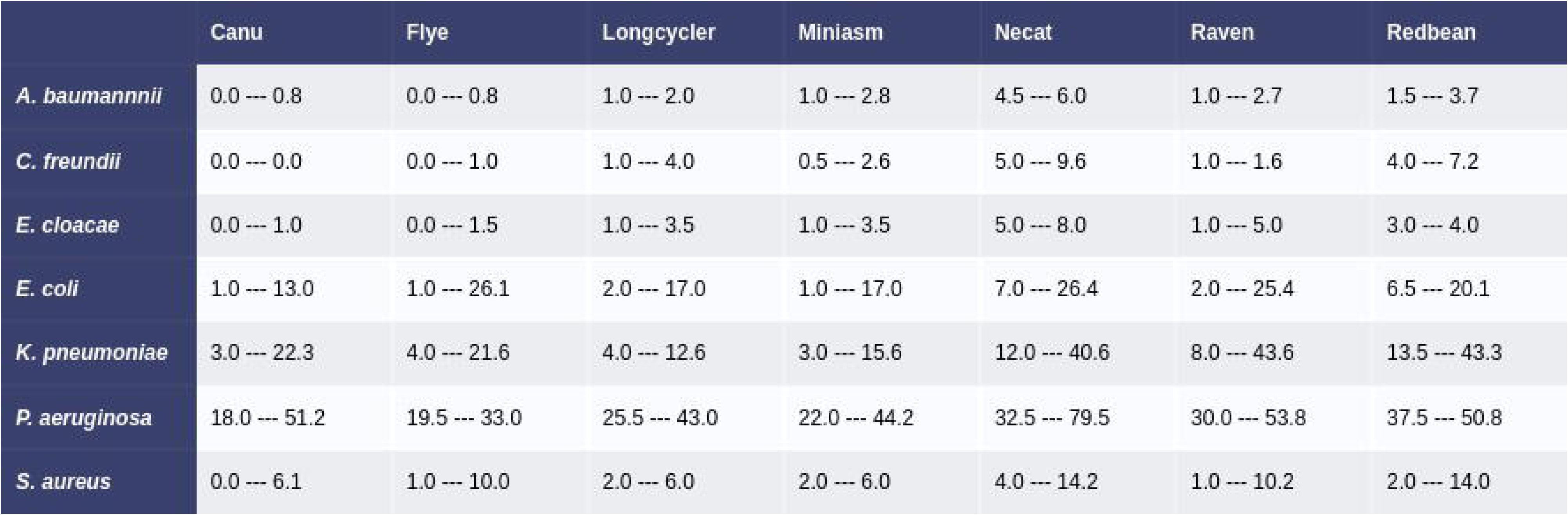
Median allele and 95 percentile wgMLST allele difference for each combination of species and *de novo* assembler.

### Detection of AMR genes and plasmid replicons in long-read sequencing data

Next, we compared the detection of AMR genes and plasmid replicons using both long-read and short-read sequencing platforms. The sensitivity and specificity of AMR gene identification was assessed. The specificity of AMR gene calling for Redbean ranged from 68.2 to 100 (median 87.2) and performed worst of all assemblers tested (Table 3A). Canu had the highest specificity in AMR gene calling (99.7%), followed by Flye (99.6%) and Longcycler (98.6%). Even though Canu seemed to perform best, multiple copies of the same AMR gene were detected (Supplementary Figure 1), suggesting *de novo* assembly artifacts. The median number of AMR genes for all species together was 16 for Canu assemblies versus 11 to 13 for all other assemblers (Supplementary Figure 1). The effect of this phenomenon was seen in the median genome size over the entire dataset (5.6-Mbp Canu versus 5.2-Mbp for all other long read assemblers, Supplementary Figure 2). Furthermore, the number of replicons was at least double for Canu assemblies versus any other assembler (Supplementary Figure 3). The unweighted average sensitivity for AMR gene calling was 98.1% independent of which *de novo* assembler was used and seemed to be a species-specific effect (Table 3). Only for Flye and Canu assemblers all carbapenemase genes were correctly detected within the MDRO tested (Supplementary Table 3). The specificity of plasmid replicon detection was best for Canu assembled long-read data (Table 3B), but as mentioned previously, was hampered by the multi-copy assembly artifact issue (Supplementary Figure 3). Next to Canu, Flye performed excellent with an unweighted average specificity of 98.5% over all species (Table 3B). Identifying the correct plasmid replicon was excellent and did not seem to differ much among assemblers as the sensitivity was between 98.9% (Canu) and 99.9% (Flye).

**Table 3.**
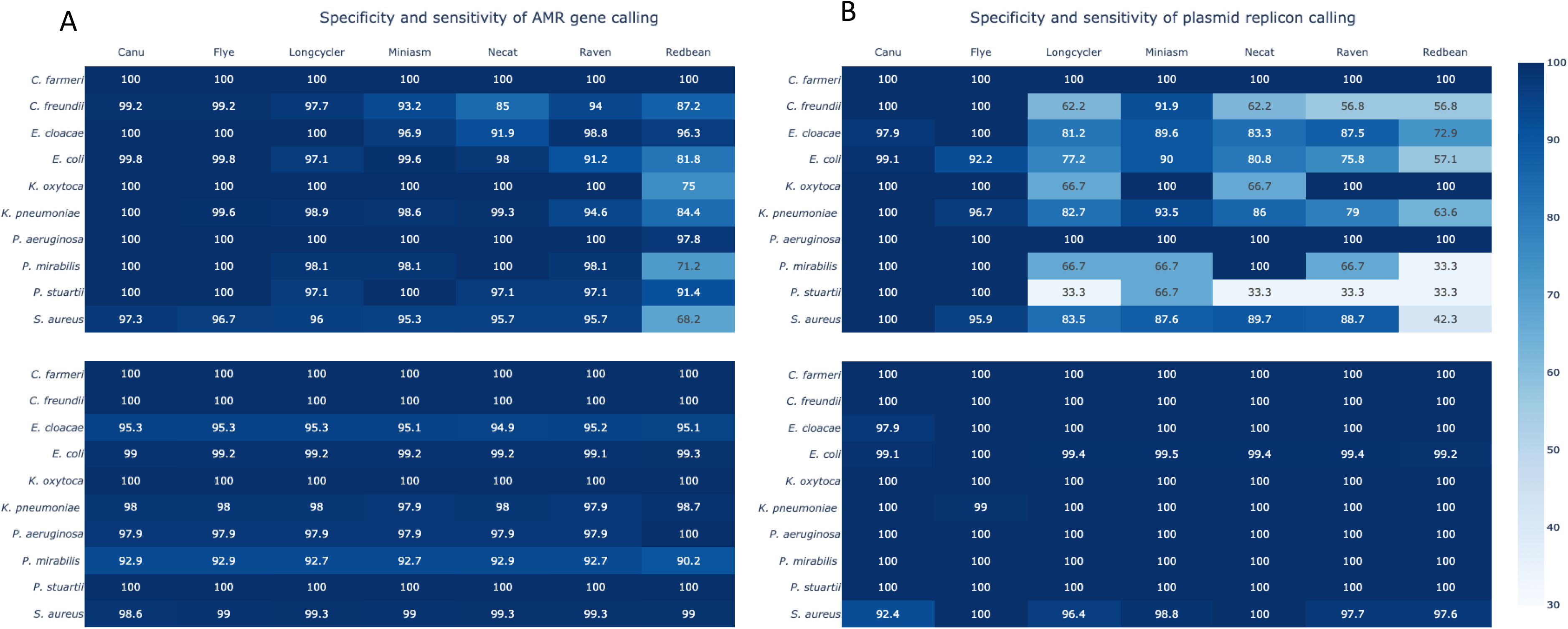
A) Percentage sensitivity (lower panel) and specificity (upper panel) of AMR gene identification for each species and assembler tested in this study. B) Percentage sensitivity (lower panel) and specificity (upper panel) of plasmid replicon detection for each species and assembler were tested in this study. Only Rerio as basecalling and long-read sequenced isolates with a coverage higher than 40x coverage were used.

### Long-read sequencing enables analysis of an MRSA outbreak

There was an impetigo-associated MRSA outbreak with MLVA-type MT4627 in the Mid-East of the Netherlands in 2019 (Figure 4A + B) [38]. The number of MRSA MT4627 isolates in the South-West region increased from 14 (in 2021) and 28 (in 2022) to 40 MRSA MT4627 isolates in 2023 and were obtained from 40 persons (Figure 4A). Characterization of a subset of the 2023 MRSA outbreak isolates using both long-read (*n*= 16) and short-read (*n*=16) sequencing-based wgMLST analysis revealed genetic clustering (≥2 isolates differing ≤15 wgMLST alleles) with 2019 outbreak and isolates from 2020, 2021, 2022 in the minimum spanning tree (Figure 4B). Long-read sequenced MRSA outbreak isolates were in close proximity of their short-read sequenced counterparts (Figure 4B). MLST, MLVA and *in silico* MLVA (iMLVA) analyses revealed that MLST sequence type and iMLVA type could be retrieved from long-read sequencing data with >40x coverage and were concordant with fragment size-based *in vitro* MLVA types. The outbreak MRSA isolates were from MLST ST121, clonal complex (CC) CC121, MLVA type MT4627 and iMLVA type MT4627. A subset of the MRSA isolates from 2023 have diversified over time. Long-read sequencing also yielded identical AMR genes *aac(6’)-aph(2’’)*, *mecA, fusC, blaZ,* and *dfrG,* plasmid replicons, and virulence genes, when compared to short-read sequencing (Figure 4C, Supplementary Table 4). The plasmid-borne *ermC* gene was lacking three long-read sequenced isolates. Long-read sequencing outperformed detection of sortase B substrate genes encoding microbial surface components recognizing adhesive matrix molecules (MSCRAMM) such as *clfB, sdrC, sdrD,* and *sdrE* (Figure 4C).

**Figure 4.**
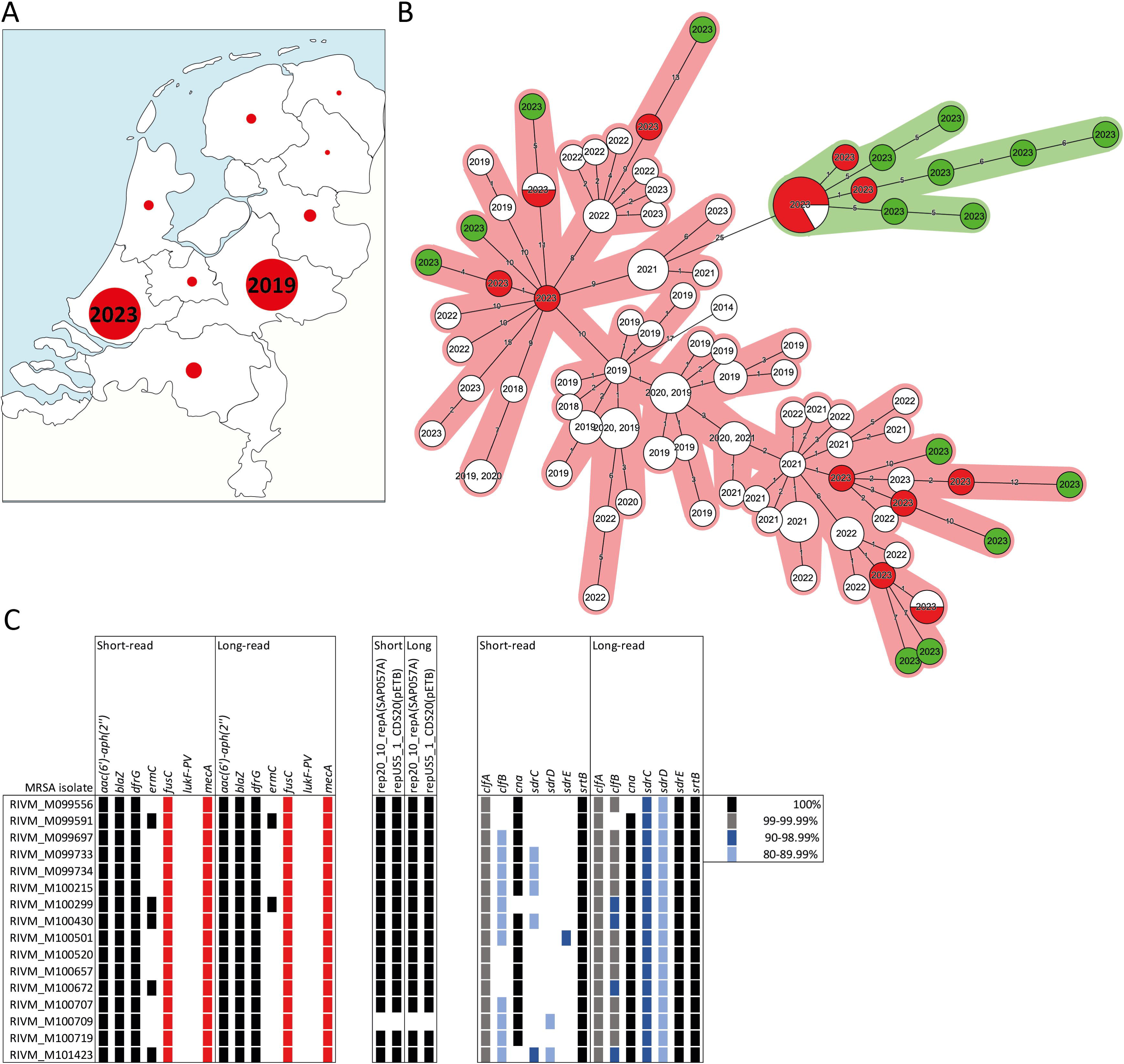
Long-read sequencing-based analyses of an impetigo-associated MRSA outbreak in the Netherlands 2023. **A)** Geographic localization of persons with an MT4627 MRSA in the Netherlands. The initial outbreak in 2019 has been described previously [38]. After this outbreak, this type mainly occurred in the province of Zuid-Holland, The Netherlands, towards the end of 2023. **B)** Minimum spanning tree of MT4627 MRSA analyzed by wgMLST. In total 16 isolates from 2023 were long-read sequenced to 40x coverage and included in the analysis (green, Table 1). Of these 16 isolates, the short-read counterpart was included in the figure (red). A wgMLST cluster cut-off of 15 was used. Halo’s indicate isolates varying ≤ 15 wgMLST alleles. **C)** Comparison of short-read and long-read sequenced resistomes, plasmid replicons, and genes encoding microbial surface components recognizing adhesive matrix molecules (MSCRAMM) of the outbreak-associated cluster isolates.

## Discussion

We demonstrate that automated DNA extraction followed by Nanopore R10.4.1 long-read sequencing is a reliable and cost-effective method for the molecular typing (iMLVA, MLST, wgMLST), AMR gene, plasmid replicon, virulence gene identification, and outbreak detection of MDRO and can therefore be applied for genomic surveillance. A sufficient sequencing depth is required, and in this study a minimum of 40x sequencing depth was used. Previous studies using Illumina short-read sequencing used 30x coverage as a minimum [41,42]. Older Nanopore flowcell generations had higher error rates of raw Nanopore reads compared to Illumina raw reads and may explain that still a higher coverage is required for R10.4.1 with current models. Long-read based wgMLST seemed to be more efficient for species with a small chromosome e.g., *S. aureus* than for those with larger chromosomes like *P. aeruginosa*. Unique signatures in *P. aeruginosa* data and or a lack of representation in the training set for the models is one explanation for poor performance. Another possibility is that even higher sequencing depth is required for bacteria with large chromosomes to obtain sufficient good quality reads. To overcome this problem, one could reload the DNA library prep during a long-read sequencing run. Alternatively, we recently performed a Promethion run with 96 MDRO which boosted coverage from 52x to 669x, with median coverage of 220x (data not shown). Two different basecalling models were tested using Dorado duplex mode with and without Medaka polishing and Rerio, this model has an increased performance for highly methylated DNA motifs. Overall, Rerio performed best for wgMLST allele calling using in-house wgMLST schemes. Research models such as Rerio are continuously updated and if proven successful, implemented into the normal workflow as has been the case for this research model where the same training set was used and now included in dna_r10.4.1_e8.2_400bps_sup@v4.3.0. Although these updates provide a constant improvement of the ability to perform molecular characterization of bacterial isolates, it imposes a problem for laboratories using Nanopore long-read sequencing under quality assurance systems. The implementation of guidelines such as ISO15189 for national reference and medical microbiology laboratories, or ISO23418 for food-borne pathogen sequencing requires the validation of every new basecalling model before implementation and is slowing down the introduction of improved methods.

For *de novo* assembly, Canu was unsuitable for AMR gene detection as it resulted in the erroneous detection of multiple copies of the same gene. This is likely due to the inability of this algorithm to properly overlap contig ends. Although Trycycler [43] is among the best long-read only *de novo* assembler in the community, this tool was not used as it requires multiple manual curation steps in each assembly, thereby hampering the ability for automated high-throughput bacterial assemblies. The use of Miniasm or Longcycler (Unicycler supplied with long-reads only) resulted in the lowest number of different wgMLST alleles compared to their Illumina short-read sequenced counterparts. Besides *Pseudomonas*, all other species tested had only a few discrepant wgMLST allele calls (0 to 12 alleles). This was on the same level of variation in a recent multi-center study where Enterobacterales, Enterococci and Staphylococci isolates were sent to participating laboratories for short-read sequencing and subsequent molecular typing analysis [41,42]. For identification of AMR genes and plasmid replicons Flye performed better than Longcycler and Miniasm, even though these two assemblers were best for wgMLST allele calling. Overall, genotyping was excellent and on par with other studies investigating the inter-laboratory reproducibility of Illumina short-read sequencing-based genotyping, where they found >99% performance [41,44]. Notably, long-read sequencing-based *de novo* assembly methods can better discriminate multi-copy AMR genes in mobile genetic elements such as plasmids, as these regions are impossible to assemble with short-read sequencing. Therefore, the evaluation of the specificity for AMR genes and replicon identification may not be the best metric to evaluate the performance of long-read sequencing methods when using Illumina short-read sequencing data as golden standard. Additionally, short-read assembly is unable to truly identify multi-copy AMR genes. However, no other reference is available. Finally, it should be noted, genotype to phenotype is still difficult to infer, and large discrepancies have been observed among methods used in a multi-centre study investigating this challenge [45]. Furthermore, Nanopore long-read sequencing was superior over Illumina short-read sequencing of genes encoding MSCRAMMs. MRSCRAMMs are known to harbor multiple repetitive domains and are implicated in binding to collagen, fibrinogen, cytokeratin components of the extracellular matrix [46].

In conclusion, for laboratories wanting to implement Nanopore long-read sequencing of MDRO we therefore recommend using a minimum 40x coverage, Rerio basecalling and Miniasm or Longcycler as *de novo* assembler for molecular typing and outbreak detection for genomic surveillance. For the best performing AMR gene and plasmid replicon detection we recommend using Flye instead of Canu, Miniasm or Longcycler, as Canu generated assembly artifacts and Miniasm and Longcycler did not perform as good as Flye on sensitivity and specificity. Future studies are needed to optimize the performance of Nanopore long-read sequencing for *P. aeruginosa*. The use of long-read sequencing can provide additional valuable insights into virulence determinants, resistance plasmids and resistance gene copy number of MDRO. This may help to inform and guide effective control measures for MDRO which were previously not possible using short-read sequencing. Importantly, the relatively low purchase and implementation costs of long-read sequencing, coupled with the low price per isolate and rapid library preparation not only enables genomic surveillance and outbreak analysis but extends its applicability to resource-constrained settings and low-income countries world-wide.

## Ethical statement

Ethical approval was not required for the present study, since it is based on genomic surveillance data only. Samples from which the isolates were cultured, were all collected as part of routine health care.

## Acknowledgements

We thank all the members of the Dutch CPE and MRSA surveillance study Groups and the Dutch medical microbiology laboratories for submitting MDRO isolates to the RIVM for the national CPE/MRSA surveillance program. We thank Dr. Romy D. Zwittink, Dr. Rob Mariman and Dr. Daan W. Notermans for critical reading of the manuscript. Members of the Dutch CPE and MRSA Surveillance Study Groups:

- A.L.E. van Arkel, ADRZ medisch centrum, Department of Medical Microbiology, Goes
- M.A. Leversteijn-van Hall, Alrijne Hospital, Department of Medical Microbiology, Leiden
- W. van den Bijllaardt, Amphia Hospital, Microvida Laboratory for Microbiology, Breda
- R. van Mansfeld, Amsterdam UMC - location AMC, Department of Medical Microbiology and Infection Prevention, Amsterdam
- K. van Dijk, Amsterdam UMC - location Vumc, Department of Medical Microbiology and Infection Control, Amsterdam
- B. Zwart, Atalmedial, Department of Medical Microbiology, Amsterdam
- B.M.W. Diederen, Bravis Hospital/ZorgSaam Hospital Zeeuws-Vlaanderen, Department of Medical Microbiology, Roosendaal/Terneuzen
- H. Berkhout, Canisius Wilhelmina Hospital, Department of Medical Microbiology and Infectious Diseases, Nijmegen
- D.W. Notermans, Centre for Infectious Disease Control, National Institute for Public Health and the Environment, Bilthoven
- A. Ott, Certe, Department of Medical Microbiology Groningen & Drenthe, Groningen
- K. Waar, Certe, Department of Medical Microbiology Friesland & Noordoostpolder, Leeuwarden
- W. Ang, Comicro, Department of Medical Microbiology, Hoorn
- J. da Silva, Deventer Hospital, Department of Medical Microbiology, Deventer
- A.L.M. Vlek, Diakonessenhuis Utrecht, Department of Medical Microbiology and Immunology, Utrecht
- A.G.M. Buiting, Elisabeth-TweeSteden (ETZ) Hospital, Department of Medical Microbiology and Immunology, Tilburg
- L.G.M. Bode, Erasmus University Medical Center, Department of Medical Microbiology and Infectious Diseases, Rotterdam
- Jansz, Eurofins PAMM, Department of Medical Microbiology, Veldhoven
- S. Paltansing, Franciscus Gasthuis & Vlietland, Department of Medical Microbiology and Infection Control, Rotterdam
- A.J. van Griethuysen, Gelderse Vallei Hospital, Department of Medical Microbiology, Ede
- J.R. Lo Ten Foe, Gelre Hospital, Department of Medical Microbiology and Infection Control, Apeldoorn
- M.J.C.A. van Trijp, Groene Hart Ziekenhuis, Department of Medical Microbiology and Infection Prevention, Gouda
- M. Wong, Haga Hospital, Department of Medical Microbiology, ‘s-Gravenhage
- A.E. Muller, HMC Westeinde Hospital, Department of Medical Microbiology, ‘s-Gravenhage
- M.P.M. van der Linden, IJsselland hospital, Department of Medical Microbiology, Capelle a/d IJssel
- M. van Rijn, Ikazia Hospital, Department of Medical Microbiology, Rotterdam
- S.B. Debast, Isala Hospital, Laboratory of Medical Microbiology and Infectious Diseases, Zwolle
- E. Kolwijck, Jeroen Bosch Hospital, Department of Medical Microbiology and Infection Control, ‘s-Hertogenbosch
- N. Al Naiemi, LabMicTA, Regional Laboratory of Microbiology Twente Achterhoek, Hengelo
- T. Schulin, Laurentius Hospital, Department of Medical Microbiology, Roermond
- S. Dinant, Maasstad Hospital, Department of Medical Microbiology, Rotterdam
- S.P. van Mens, Maastricht University Medical Centre, Department of Medical Microbiology, Infectious Diseases & Infection Prevention, Maastricht
- D.C. Melles, Meander Medical Center, Department of Medical Microbiology, Amersfoort
- J.W.T. Cohen Stuart, Noordwest Ziekenhuisgroep, Department of Medical Microbiology, Alkmaar
- P. Gruteke, OLVG Lab BV, Department of Medical Microbiology, Amsterdam
- A. van Dam, Public Health Service, Public Health Laboratory, Amsterdam
- Maat, Radboud University Medical Center, Department of Medical Microbiology, Nijmegen
- Maraha, Regional Laboratory for Microbiology, Department of Medical Microbiology, Dordrecht
- J.C. Sinnige, Regional Laboratory of Public Health, Department of Medical Microbiology, Haarlem
- E. van der Vorm, Reinier de Graaf Groep, Department of Medical Microbiology, Delft
- M.P.A. van Meer, Rijnstate Hospital, Laboratory for Medical Microbiology and Immunology, Velp
- M. de Graaf, Saltro Diagnostic Centre, Department of Medical Microbiology, Utrecht
- E. de Jong, Slingeland Hospital, Department of Medical Microbiology, Doetinchem
- S.J. Vainio, St Antonius Hospital, Department of Medical Microbiology and Immunology, Nieuwegein
- E. Heikens, St Jansdal Hospital, Department of Medical Microbiology, Harderwijk
- M. den Reijer, Star-shl diagnostic centre, Department of Medical Microbiology, Rotterdam
- J.W. Dorigo-Zetsma, TergooiMC, Central Bacteriology and Serology Laboratory, Hilversum
- A. Troelstra, University Medical Center Utrecht, Department of Medical Microbiology, Utrecht
- E. Bathoorn, University of Groningen, Department of Medical Microbiology, Groningen
- J. de Vries, VieCuri Medical Center, Department of Medical Microbiology, Venlo
- D.W. van Dam, Zuyderland Medical Centre, Department of Medical Microbiology and Infection Control, Sittard-Geleen
- E.I.G.B. de Brauwer, Zuyderland Medical Centre, Department of Medical Microbiology and Infection Control, Heerlen
- R. Steingrover, St. Maarten Laboratory Services, Department of Medical Microbiology, Cay Hill (St. Maarten)
- Analytical Diagnostic Center N.V. Curaçao, Department of Medical Microbiology, Willemstad (Curaçao)

## Figure legends

**Supplementary Figure 1** Number of AMR genes called per assembler and species

**Supplementary Figure 2** Genome size for each species and assembler.

**Supplementary Figure 3.** Number of plasmid replicons called for each assembler and species.

