## Supplementary figures and images for "Genomic surveillance of multidrug-resistant organisms based on long-read sequencing"

### Supplemental Figure 1

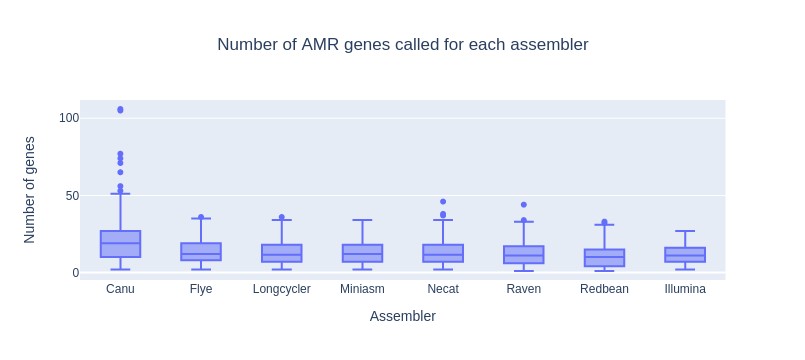

### Supplemental Figure 2

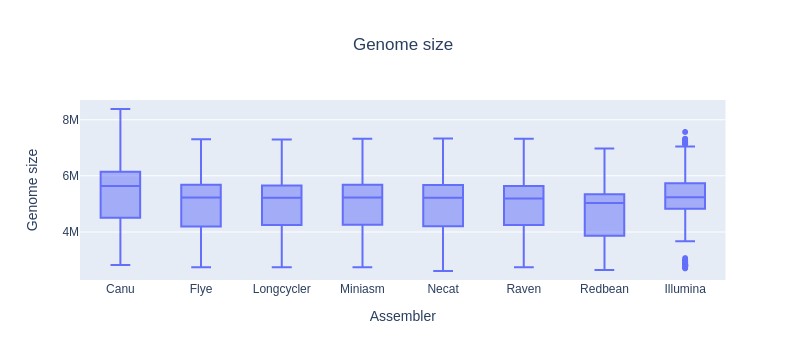

### Supplemental Figure 3

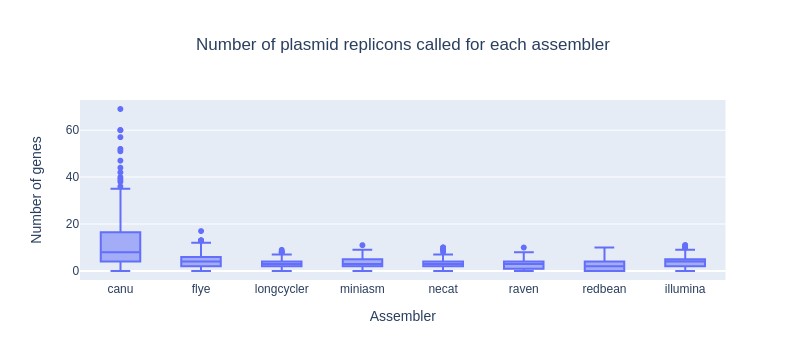

### Supplemental Table 2

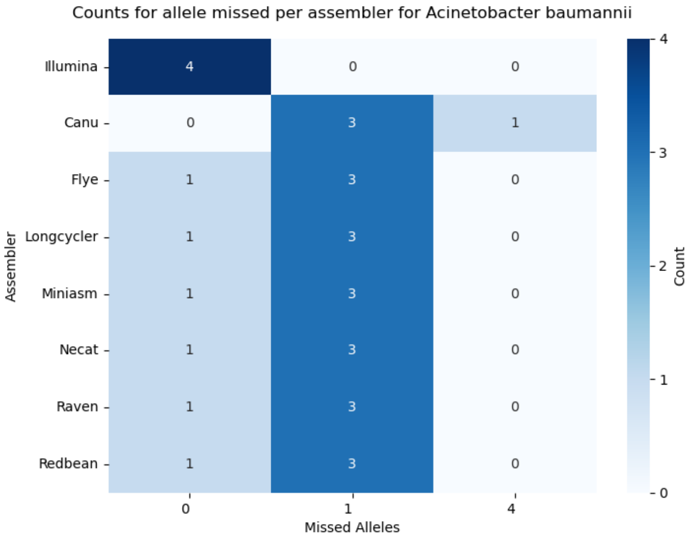

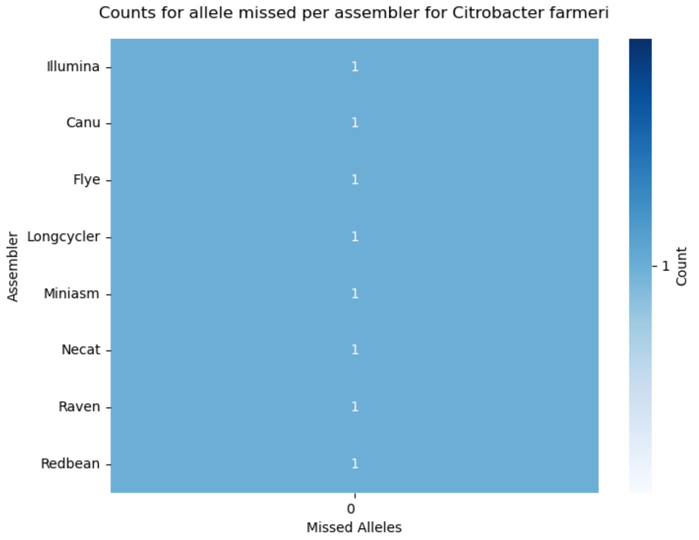

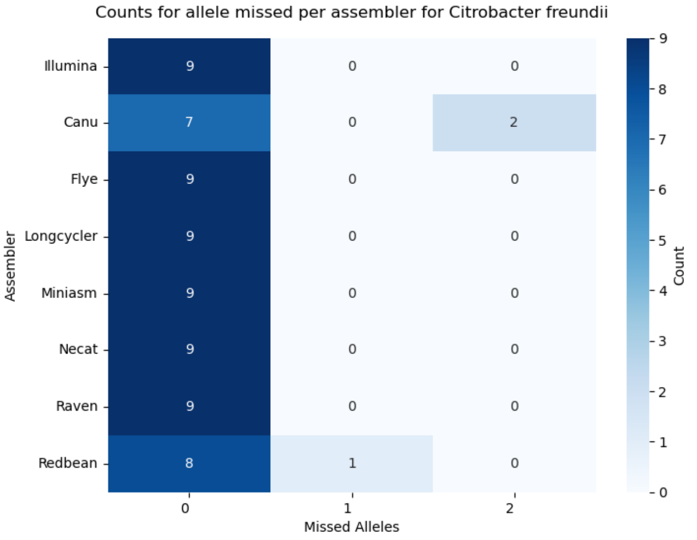

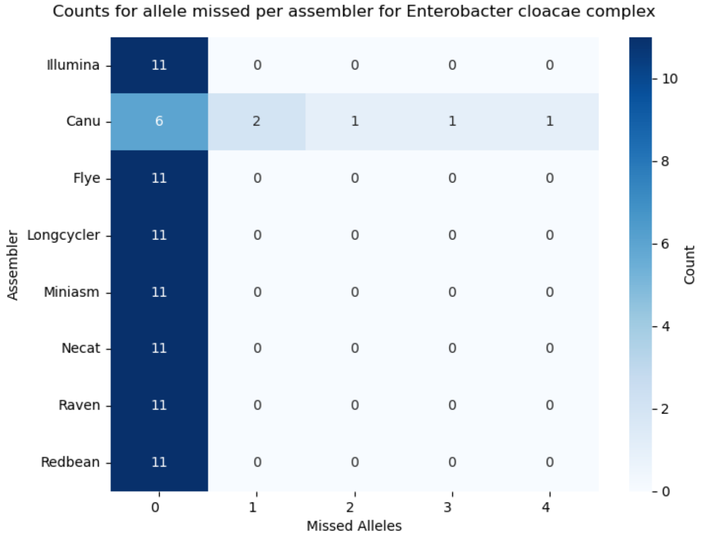

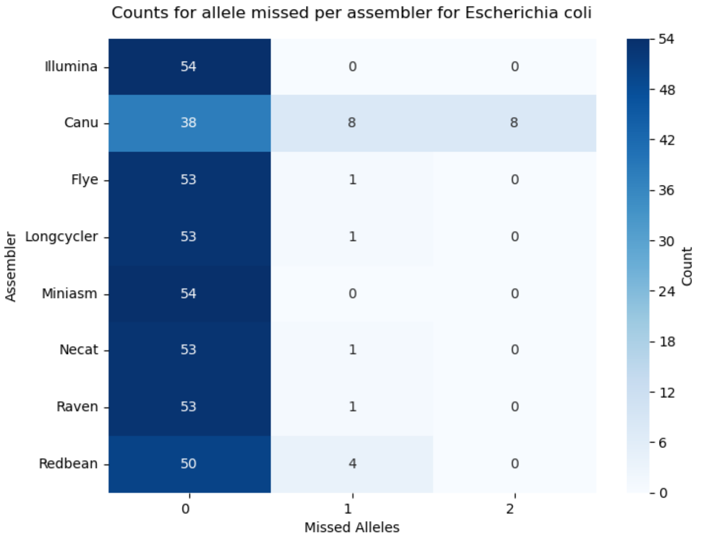

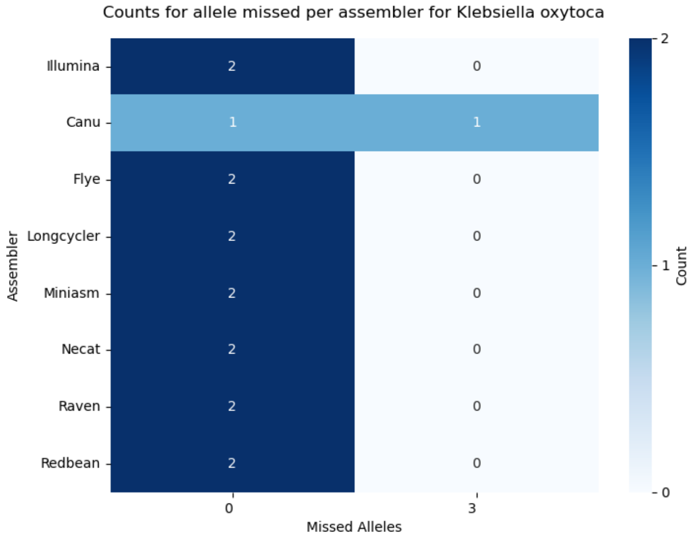

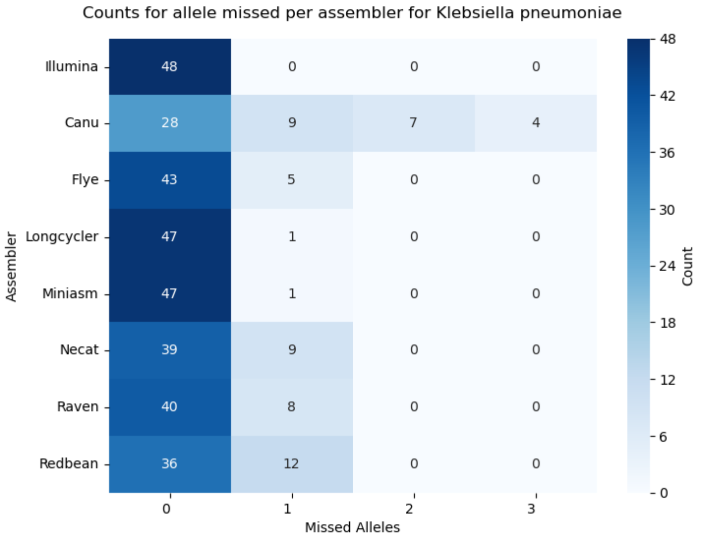

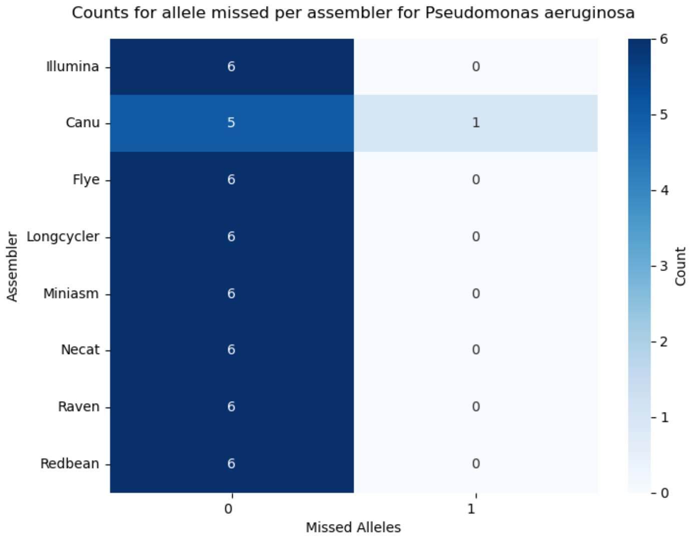

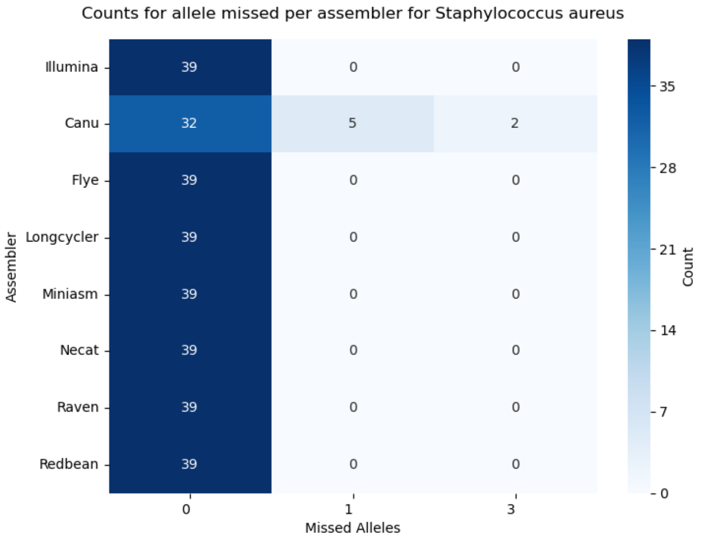
